# Public evidence on AI products for digital pathology

**DOI:** 10.1101/2024.02.05.24302334

**Authors:** Gillian A. Matthews, Clare McGenity, Daljeet Bansal, Darren Treanor

## Abstract

**Background:** Novel products applying artificial intelligence (AI)-based approaches to digital pathology images have consistently emerged onto the commercial market, touting improvements in diagnostic accuracy, workflow efficiency, and treatment selection. However, publicly available information on these products can be variable, with few sources to obtain independent evidence.

**Methods:** Our objective was to identify and assess the public evidence on AI-based products for digital pathology. We compared key features of products on the European Economic Area/Great Britain (EEA/GB) markets, including their regulatory approval, intended use, and published validation studies. We included products that used haematoxylin and eosin (H&E)-stained tissue images as input, applied an AI-based method to support image interpretation, and received regulatory approval by September 2023.

**Results:** We identified 26 AI-based products that met our inclusion criteria. The majority (73%) were focused on breast pathology or uropathology, and their primary function was tumour or feature detection. Of the 26 products, 24 had received regulatory approval via the self-certification route as General *in vitro* diagnostic (IVD) medical devices, which does not require independent review by a conformity assessment body. Furthermore, only 10 of the products (38%) were associated with peer-reviewed scientific publications describing their development and internal validation, while 11 products (42%) had peer-reviewed publications describing external validation (i.e., testing on data from a source distinct to that used in development).

**Conclusions:** The availability of public information on new products for digital pathology is struggling to keep up with the rapid pace of development. To support transparency, we gathered available public evidence on regulatory-approved AI products into an online register: https://resources.npic.uk/AI/ProductRegister. We anticipate this will provide an accessible resource on novel devices and support decisions on which products could bring benefit to patients.

## Introduction

The increasing adoption of digital pathology, combined with the expansion of machine learning-based methodologies, has opened the door for novel products that generate predictions from the morphometric features within digital images. These artificial intelligence (AI)-based technologies have the potential to support the pathology workflow by improving diagnostic accuracy, assisting in case triage, and guiding treatment selection, which could alleviate pressure on pathology services, reduce costs, and improve patient outcomes. Realising these potential benefits, however, relies upon clinical deployment of well-validated algorithms, which have undergone thorough performance and usability testing.

High standards are expected of any medical device to be used in clinical practice, including software. Many organisations worldwide provide guidance on digital technology standards to be met for clinical adoption. In the UK, for example, the requirements for clinical performance, safety, security, and cost-effectiveness are defined in documentation including: the *Evidence Standards Framework for Digital Health Technologies* from the National Institute for Health and Care Excellence (NICE)^1,2^, the *Digital Technology Assessment Criteria* from NHSX^3^, and the *Guide to Good Practice for Digital and Data-Driven Health Technologies* from the Department of Health and Social Care (DHSC)^4^. This guidance, alongside medical device legislation, is consistent in requiring new technologies to be thoroughly tested on representative datasets during clinical validation^1,4^. For digital pathology in particular, rigorous validation of algorithms on diverse test datasets is essential to demonstrate accuracy and generalisability, as the process of digital image creation can introduce site-specific variation through differences in glass slide preparation, staining protocols/reagents, scanner platforms, image formats, and scan quality^5–7^. However, as new AI-based products continue to appear on the market, it can be challenging to ascertain the level of testing a new product has undergone, including the source and composition of test datasets.

All novel medical devices must obtain regulatory approval prior to placement on the EEA/GB markets. This involves a conformity assessment process whereby manufacturers gather evidence to demonstrate compliance with medical device legislation. For software, this covers elements including safety and performance, risk management, and product verification and validation^8^. In the EEA, AI-based image analysis software is typically considered an *in vitro* diagnostic (IVD) medical device^9^, and is therefore subject to the *In Vitro* Diagnostic Medical Devices Regulation (2017/746) (IVDR^8^), which applied from May 2022 (Fig. 1A). This replaced older legislation (the IVD Directive 98/79/EC; IVDD)^10^, which is currently mirrored in the existing equivalent UK legislation (UK MDR 2002^11^). One crucial difference between the original IVDD/UK MDR and new IVDR is that, under IVDD, AI-based image analysis software for digital pathology was typically considered a ‘General IVD’, which is the lowest device class. This allows manufacturers to ‘self-certify’ their device as conforming with essential requirements (Fig. 1B). Contrastingly, under the new IVDR, similar software is more likely to be a higher-risk Class C device, and therefore must meet higher safety and performance standards, and requires evidence review by a conformity assessment body. At present, there is a transitional period which allows certain IVDD-compliant devices with a valid *Conformité Européenne* (CE)-mark to continue to be placed on the EEA market up to May 2027, depending on the device class (Fig. 1A). Following withdrawal from the EU, different rules now apply to the GB market^12,13^ (Fig. 1A-B). Currently, most IVDs registered with the MHRA can be placed on the GB market if they: (1) conform to the UK MDR 2002 requirements to receive the UK Conformity Assessed (UKCA) marking; or (2) if they have a valid certificate or declaration of conformity and CE-mark issued before May 2022 under the EU IVDD; or (3) if they have a valid certificate or declaration of conformity and CE-mark under the new EU IVDR^14–16^ (see Fig. 1A-B for further details). Consequently, AI-based medical devices on the EEA/GB market across this ongoing transition period may be compliant with different standards and requirements, which compels the need for independent evaluation of novel emerging products.

**Fig. 1.**
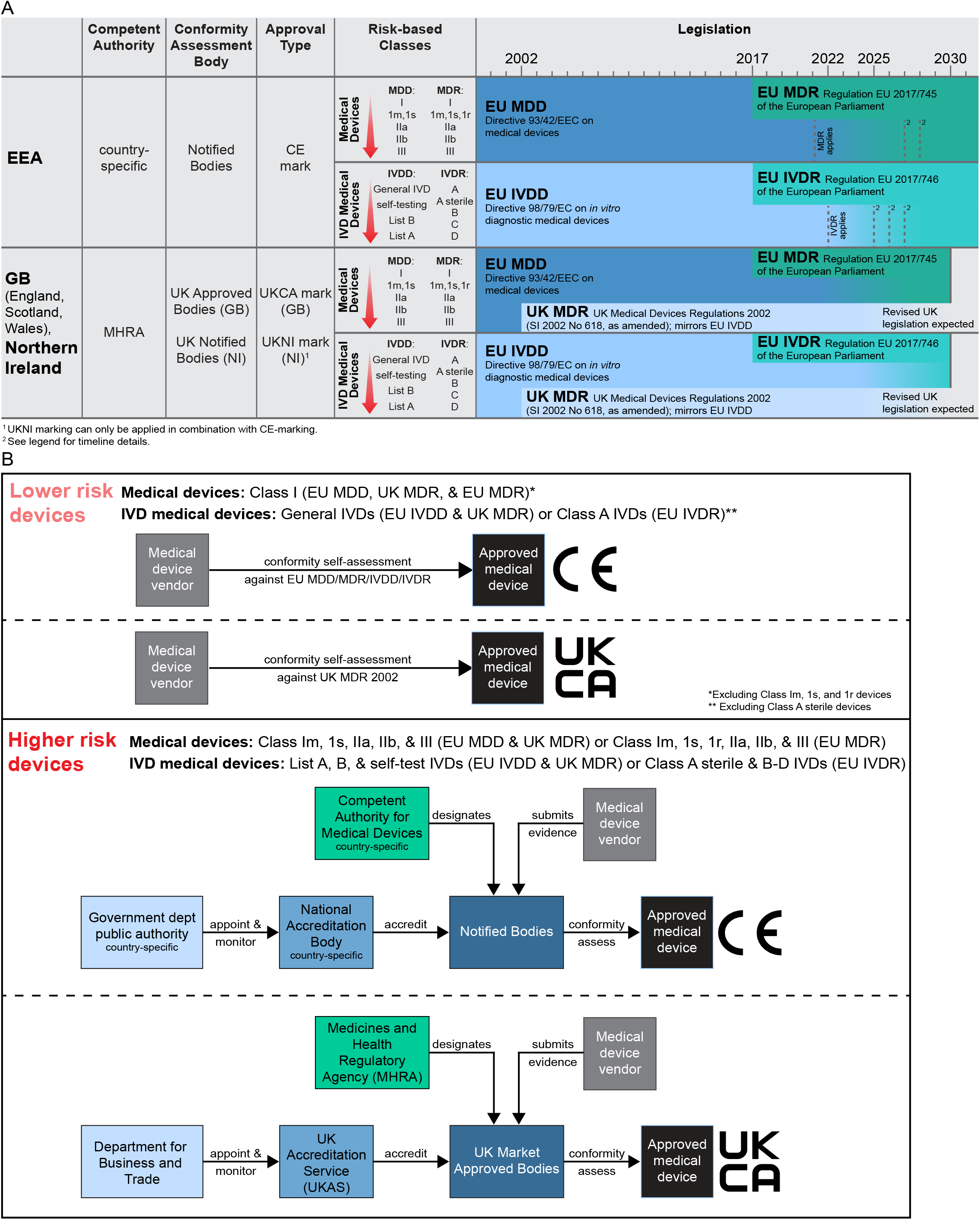
Conformity Assessment of Medical Devices. (**A**) Table comparing the relevant bodies involved in regulatory approval in the EEA and GB/Northern Ireland (NI), and the timeline for acceptance of compliant devices under different legislation. The EU MDR and EU IVDR entered into force in 2017 and became applicable in 2021 and 2022, respectively. For placement on the EEA market, there is a transitional period until December 2027 for higher risk devices (i.e., Class III and IIb implantable devices) and December 2028 for medium and lower risk devices (i.e., other Class IIb devices, Class IIa, Class Im, Is, and Ir devices). Subject to certain conditions (including an agreement in place with a notified body before certificate expiry), until those dates, certificates issued by a notified body under the old EU MDD, that were valid in May 2021, will remain valid. Additionally, Class I devices with an existing self-declaration of conformity (as of May 2021), which now require notified body involvement under the new EU MDR, can be placed on the market until December 2028^52^. Self-certified Class I devices not requiring notified body involvement under EU MDR have no transitional period and must conform with EU MDR from May 2021. Similarly, subject to certain conditions, IVDs with a valid certificate from a notified body, issued under Annex VI of the IVDD before May 2022, can be placed on the market up until May 2025^52,53^. For General IVDs with manufacturer self-declared conformity under the EU IVDD before May 2022, that now require notified body involvement under IVDR, the transition period is until May 2025 for higher risk Class D IVDs, May 2026 for Class C IVDs, and May 2027 for Class B and sterile Class A IVDs^52,53^. Non-sterile Class A IVDs, which do not require notified body involvement under EU IVDR, and new IVDs, have no transitional period and must conform with the EU IVDR from May 2022. For placement on the GB market, medical devices with a valid certificate and CE-mark compliant with the EU MDD prior to May 2021 or EU IVDD prior to May 2022, can be placed on the market up until June 2028 or June 2030, respectively, as long as the certificate remains valid. Class I medical devices or General IVDs with self-declared conformity under EU MDD/EU IVDD, that will be upclassified under the new EU MDR/EU IVDR to require notified body involvement, can also continue to be placed on the GB market, subject to certain conditions, as long as the declaration remains valid^16^. Additionally, devices with a valid CE-mark compliant with the EU MDR or EU IVDR can be placed on the GB market until June 2030^54^. Products with a UKCA mark (currently conforming to the UK MDR 2002) can also be placed on the GB market. Revised legislation is anticipated to apply beyond 2030. Under the terms of the Northern Ireland Protocol, different rules apply to devices placed on the market in Northern Ireland^14^. See MHRA timeline for full details^15^. Red arrow indicates increasing risk; Im=Class 1 measuring function, 1s=Class 1 sterile, 1r=Class 1 reusable. Conformité Européene (CE). (**B**) Summary of the process and organisations involved in conformity assessment in the EEA and GB, for different device classes, under different legislation. In the EEA, conformity assessment bodies (Notified Bodies) are accredited by country-specific National Accreditation Bodies and designated by the Competent Authority for Medical Devices. For the GB market (defined as England, Scotland, and Wales, but excluding Northern Ireland; NI), conformity assessment bodies (Approved Bodies) can only provide UKCA product marking, with UKAS being the National Accreditation Body and the MHRA the Competent Authority. The Department for Business and Trade is responsible for appointing and monitoring UKAS.

In the USA, a similar risk-based classification system is used for medical devices, with The Food and Drug Administration (FDA) being responsible for market approval^17^. Devices must conform to the regulatory controls in the Federal Food, Drug, and Cosmetic Act and the Code of Federal Regulations Title 21. The three most common routes for medical device market approval are: (1) The Premarket Notification/510(k) approval pathway, which is for medium risk devices (Class I & II) and requires vendors to prove substantial equivalence to an already-marketed device; (2) the Premarket Approval pathway, which is for high risk (Class III) and novel devices; and (3) the *De Novo* Classification Request pathway, which is for novel Class I and II devices. In addition, Class I and some Class II products can be exempt from premarket submission as long as they comply with regulatory controls laid out in the relevant legislation. To date, the only AI-based product for digital pathology to receive approval for placement on the USA market was approved in September 2021 via the *De Novo* pathway as a Class II device^18^.

Although conformity assessment does not guarantee a product will make it into clinical use, it is a crucial step in making a product available on the market and commencing further evaluation and procurement activities. However, in the EEA/GB there is currently no requirement to make the evidence provided for conformity assessment public, even in summary form^19^. The EU recently established the EU Database on Medical Devices (EUDAMED)^20^, alongside the transition to IVDR, which will provide public, searchable listings of all companies producing medical devices, as well as details of their products, certifications, approvals etc. However, this is under construction and currently contains limited information. This contrasts with the USA, where the FDA maintains databases of approved medical devices^21^, and publishes detailed decision summaries on approved devices^22^. In the UK, public evidence on medical devices can be released when products undergo a Health Technology Evaluation by NICE, which is associated with an independent, published evaluation summary. However, these evaluations are typically only conducted on select products which have received regulatory approval and have been recommended for a NICE evaluation. In radiology, NICE Guidance has so far been released for three forms of AI-based technology for medical image analysis^24^. In digital pathology, there are no AI-based products associated with NICE Guidance, however, one product has undergone a NICE MedTech Innovation Briefing^25^, which is a technology summary classed as ‘NICE Advice’ and is designed to support local decision-making rather than providing a recommendation or constituting formal NICE Guidance. Thus, for novel regulatory-approved products, it can be challenging to locate reliable public information about a product and its clinical evidence.

Maintaining transparency around novel medical devices, particularly those using AI-based methods, is crucial in promoting confidence, trust, and understanding. Details on novel technologies should be easily accessible for pathologists, patients, clinicians, commissioners, and other key stakeholders, to support informed decision-making, and promote the implementation of safe and effective technologies. In radiology, one initiative has created and maintained an index of all known AI-based products on the EU market^26^. This currently details 210 commercially available tools for radiology, with details including their regulatory approval, intended use, integration, and associated publications^26,27^. Similarly, the Royal College of Radiologists and NHS England have recently proposed to construct an open UK-based registry^28^, to document where AI-based products are in use, and to facilitate comparison of their performance. In the USA, there is also an independent database dedicated to AI-based medical devices approved by the FDA, with details on how they were evaluated^29^. However, we are not aware of any resources that currently provide a public registry of commercially available AI products on the EEA/GB market for digital pathology.

In light of these evolving initiatives, and to facilitate transparency, we sought to identify and analyse the public evidence on commercially available AI-based products on the EEA/GB market for digital pathology. Here, we describe our findings in relation to the current landscape of available products, their primary function, regulatory approvals, and clinical validation. We have compiled this information into a publicly accessible registry of products, including all available evidence relating to a product’s features and performance: https://resources.npic.uk/AI/ProductRegister. It is hoped this will provide a resource to support pathologists, patients, and commissioners in understanding the tools available for diagnostic support and their associated evidence.

## Methods

### Product eligibility criteria

Our objective was to assess the publicly available evidence on regulatory-approved AI-based products in digital pathology. We limited inclusion to products which met the following eligibility criteria:

1. Products using only H&E or haematoxylin-eosin-saffron (HES)-stained whole slide images (WSIs) of tissue as input.
2. Products using a machine learning-based approach for image analysis which augments interpretation of WSIs. Products only performing tasks relating to quality control or image management were not included.
3. Products that had received a CE- or UKCA-mark for placement on the EEA/GB market (before 30/9/2023).

Products that did not meet the above criteria were excluded from further analyses. Searches and data collection were completed between 15/7/2023 and 01/10/2023.

### Product identification

To identify products for inclusion, companies that operate on the digital pathology marketplace were located by performing searches of various sources. This included:

- Advertisements or articles in pathology publications, magazines, podcasts, including The Pathologist, Pathology News, Digital Pathology Today, Pathology in Practice, AI Magazine, and Imaging Technology News.
- List of exhibitors, sponsors, and presenters at major Pathology conferences:
  ○ Digital Pathology and AI Congress (EU 2022, 2023; Asia 2023, USA 2023)
  ○ European Congress of Pathology (2022, 2023)
  ○ European Congress on Digital Pathology (2022, 2023)
  ○ IBMS Congress (2023)
  ○ Pathology Visions (2020, 2021, 2022, 2023)
  ○ Association for Pathology Informatics (Summit 2023; Digital Pathology AI Workshop 2023)
  ○ World Digital Pathology & AI UCGCongress (2023)
  ○ College of American Pathologists Annual Meeting (2023)
  ○ Next Gen Digital Pathology Conference (2023)
- List of funding award winners developing AI for digital pathology:
  ○ NHSX AI Award (2020-2023)
  ○ NIHR-funded studies (search for ‘artificial intelligence AND pathology’)
  ○ UKRI-funded studies (Technology Missions Fund, 2023; SBRI Healthcare 2018, 2022, 2023;
  ○ Biomedical Catalyst 2020)
  ○ Innovate UK funded studies (since 2004, search for ‘pathology’)
- List of contributors to Grand Challenges in Pathology: Results and leader boards where available.
- PathPixel list of AI vendors: https://www.pathpixel.net/ai-solutions
- EUDAMED database (final search performed on 15/8/2023):
  ○ nomenclature code W0202050392: Laboratory instruments for microscopic investigations - IVD medical device software.
  ○ nomenclature code W0202050382: Laboratory instruments for microscopic investigations – software accessories.
  ○ nomenclature code W0202059092: Various processing instruments for histology/cytology – IVD medical device software.
  ○ nomenclature code W02029092: Various haematology/histology/cytology instruments – IVD medical device software.
  ○ nomenclature code W02020692: Rapid test haematology/histology/cytology instruments – IVD medical device software.
  ○ nomenclature code W02079092: Various general purpose IVD instruments – IVD medical device software.
- MHRA Public Access Registration Database (PARD; final search performed on 15/8/2023):
  ○ GMDN Code: 58721; Histology/cytology/microbiology image-analysis interpretive software IVD; Medical Risk Classification A, C, and IVD General.
  ○ GMDN Code 43472; Laboratory instrument/analyser application software IVD; Class A and IVD General.
- FDA databases:
  ○ AccessGUDID (Global Unique Device Identification Database).
  ○ Devices@FDA.
- Industry partners of the five UKRI-funded Centres of Excellence in Digital Imaging and Artificial Intelligence.

In addition, other newsletters, press releases, and general news reports were also scrutinised for any novel developments. For every company identified in the initial search, the company’s public website was used to ascertain whether they developed AI-based products that supported the interpretation of WSIs. If it could not be determined whether a product met all eligibility criteria from the website (and links therein), additional sources were searched (e.g., internet searches for news items or press releases relating to the company). Information accessed via the company’s website and additional searches were used to determine if a product met all criteria for inclusion.

### Data collection on eligible products

For eligible products, public information was collected on the company and the product (Table S1). Information relating to the company was sourced from the company’s LinkedIn page (for employee number) and their website. Information relating to the product was sourced from the company’s website, internet searches for news items or press releases, PubMed, the Sectra Amplifier Marketplace, and three databases of medical device manufacturers (EUDAMED, PARD, and FDA). Products forming part of a ‘suite’ marketed by vendors were considered to be separate products for the purpose of this analysis, as they perform distinct functions and were often separately submitted for regulatory approval.

Scientific publications relating to the products were identified by manually searching many of the sources described above (i.e., company website, news sites, Sectra Amplifier Marketplace). In addition, searches were performed on PubMed, bioRxiv, arXiv, medRxiv, and Google Scholar, by including the company name and product disease/tissue focus. Publications were included if they were an original study, written in English, and described the development or testing of the algorithm on relevant human data. Preprints were also included (where noted).

For each publication, data was extracted under a specified list of fields (Table S2). This included, were available, information relating to the public accessibility of the publication, the independence from vendors, whether it described an internal or external clinical validation study, and details on the study itself (e.g., the datasets used, the source of data, scanner platforms included, and algorithm performance). A publication was determined to be open access if it was possible to access the full publication materials without a journal or institutional subscription or an additional payment. To determine whether a publication was independent of vendors, author affiliations and funding source were scrutinised. Publications were defined as independent if authors were not employees of, or shareholders in, the company and the study was not funded by the company. To classify the type of study described (internal or external validation), internal validation studies were defined as those using test data from the same institution(s) as algorithm training, whereas external validation studies were defined as those using test data from a different institution(s) to algorithm training. In cases where multiple products were described within the same publication, these were considered as separate studies when extracting information relating to the datasets used for training/testing and performance metrics. Thus, there are more studies described than publications (see Table S2). When examining the datasets used for training/testing, a dataset ‘source’ was defined as a distinct site from which samples originated (e.g., lab or institution), even if those samples were then processed or scanned at a different location. Performance metrics were extracted that related to the standalone performance of the product and/or the performance when the product was used by pathologists. Slide or case-level performance was reported wherever possible, and any analyses of impact on workflow were noted when this information was available.

Information was extracted from publications by two reviewers, and the consensus between the two reviewers was taken for analysis. It was not always possible to verify how the algorithm described in a publication related to the final approved product, i.e., the publication may have described work on an earlier version of the algorithm which has since been modified or updated to generate the final product. Data extracted on each product and each publication was collated in spreadsheets, data was analysed in Microsoft Excel (Microsoft Corporation, NM, USA), and figures generated in Adobe Illustrator (Adobe Systems Incorporated, CA, USA).

### Public Product Register

The information compiled on eligible AI-based products with regulatory approval has been made available online at: https://resources.npic.uk/AI/ProductRegister. We intend to maintain and update the product register at regular intervals, and include information provided by suppliers where this is made available. In future, the database will also be updated to include other digital pathology AI-based products, including those for special stains and immunohistochemistry (IHC).

## Results

### Product identification

Initial searches to identify eligible AI-based products located 400 companies producing digital pathology-related products (Fig. 2A). Company offerings were reviewed, which revealed 60 companies developing AI-based image analysis software and 115 products that performed analysis on H&E-stained tissue images. Of these, 26 products from 15 unique companies had received regulatory approval for placement on the EEA/GB market and met all eligibility criteria for inclusion (Fig. 2B). A summary of the details gathered in relation to all 26 eligible products is shown in Table S1.

**Fig. 2.**
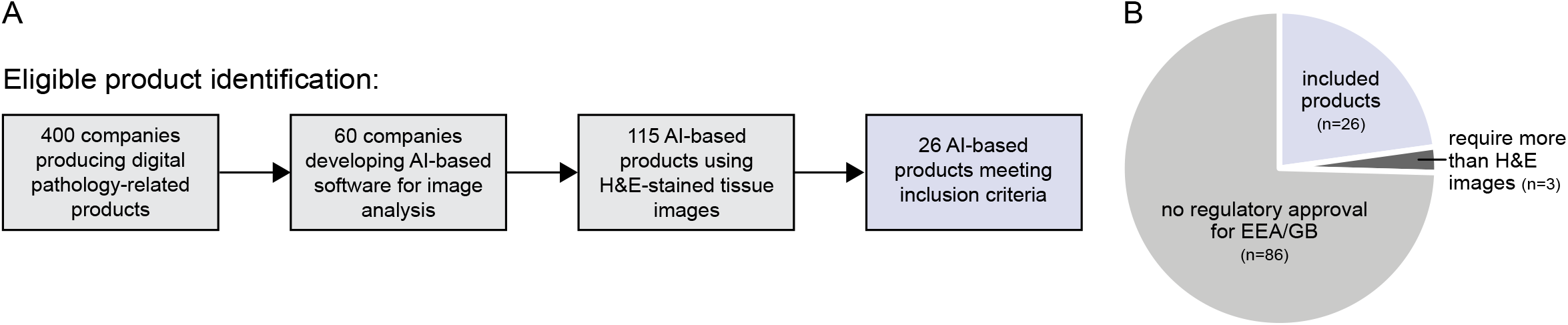
Product identification. (**A**) Flow diagram indicating the number of companies and products identified during searches, including the total that met all inclusion criteria. (**B**) Pie chart showing the proportion of tools that met criteria for further analysis.

### Key product features

The 15 companies producing the 26 eligible products were headquartered across 11 different countries, and approximately half of these companies (8/15) had <50 employees (Fig. 3A; Table S1). Of the 26 products, 24 had achieved regulatory approval for the EEA market as ‘General IVDs’ under IVDD, and two as Class C devices under IVDR (Fig. 3B). Nine of the 26 products had also received a UKCA mark, and one had received FDA approval.

**Fig. 3.**
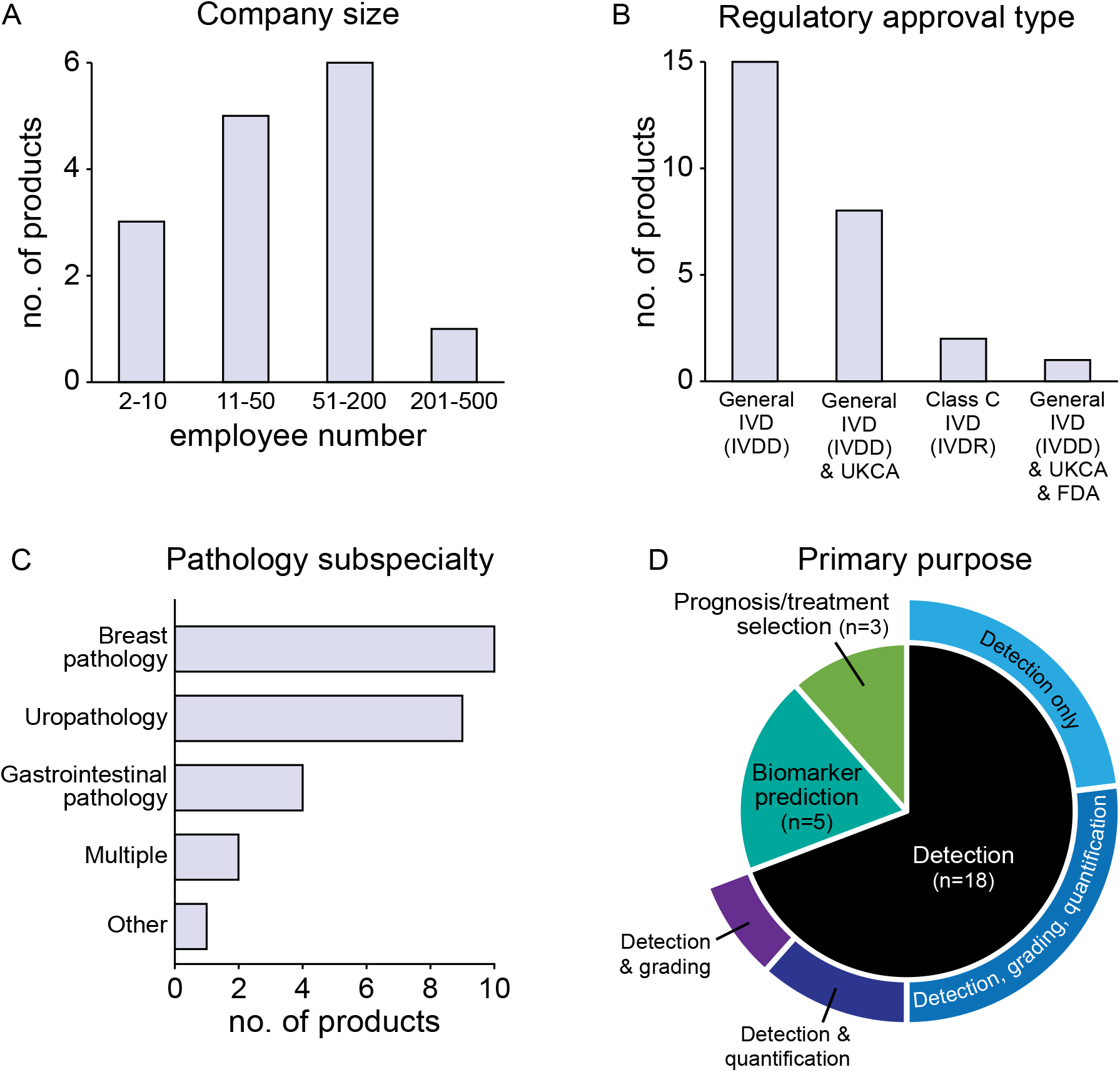
Key features of eligible products. Graphs showing (**A**) the size of companies producing eligible AI-based digital pathology products, (**B**) the type of regulatory approval received, (**C**) the pathology subspecialty, and (**D**) the primary purpose of each the product. *In Vitro* Diagnostic Device (IVD); Directive 98/79/EC on *In Vitro* Diagnostic Medical Devices (IVDD); Regulation (EU) 2017/746 on *In Vitro* Diagnostic Medical Devices (IVDR); UK Conformity Assessed (UKCA); United States Food and Drug Administration (FDA).

Both the EU and UK aim to provide transparent listings of companies producing commercially available medical devices, through EUDAMED^20^ and the MHRA’s Public Access Registration Database (PARD^30^), respectively. EUDAMED intends to provide details of registered companies, medical devices, product classification, certificates, and clinical investigation. However, it currently only contains limited information and is expected to be completed in 2024. The MHRA’s PARD is updated weekly and contains listings of registered manufacturers, alongside the category and classification of their medical devices, but does not list the specific product name. Notably, the MHRA requires all medical devices to be registered with the MHRA before placement on the UK market (including NI)^14^. Performing a search of these databases revealed that 11 of the 15 companies (producing 22 of the 26 CE-marked tools) were currently registered as ‘actors’ (typically manufacturers or authorised representatives) on the EUDAMED database, but only one had an AI-based medical device listed. Furthermore, of the nine UKCA-marked products, all companies were listed as a manufacturer on MHRA’s PARD database. However, only 17 of the 26 CE-marked products had manufacturers listed on the MHRA database. In addition, the one tool that had received FDA approval was located on their medical device database^21^ and had an associated classification order and decision summary available.

The majority (73%) of products that met inclusion criteria were focused on breast pathology (38%, 10/26 products) and uropathology (35%, 9/26 products; Fig. 3C). The remaining seven products interpreted images relating to gastrointestinal pathology (15%, 4/26 products) or multiple/other pathology subspecialities. The primary purpose of the products was tumour or feature detection (69%, 18/26 products) with six performing detection only, seven performing detection with grading & quantification, and five performing detection with either grading or quantification (Fig. 3D). The purpose of the remaining tools was biomarker prediction (19%, 5/26 products) and prognosis/treatment selection (12%, 3/26 products).

### Scientific publications associated with eligible products

For all eligible products, scientific publications describing their development or testing were identified. Specified data was extracted from each publication, including the composition of training and testing datasets, the algorithm performance, and whether the publication was open access and/or independent of the vendors (see *Methods* for criteria used to make determinations). All data extracted from each publication is shown in Table S2. Of the 26 products, 15 were associated with scientific publications (Fig. 4A). This included 23 publications (18 peer-reviewed and five preprints), three of which described internal validation studies, six of which described external validation studies, and 14 of which described both types of study (Fig. 4B). Therefore, of the 26 regulatory approved products, 50% (13/26) were associated with published details of an internal validation study, and 38% (10/26) were associated with *peer-reviewed* evidence of an internal validation study. Similarly, 54% of products (14/26) were associated with published details of an external validation study, and 42% (11/26) were associated with peer-reviewed evidence of an external validation study. In addition, 87% of the available scientific publications (20/23) were open access, but only 17% (4/23) were independent of vendors (i.e., the company and its employees neither authored nor funded the study; Fig. 4B). To appreciate the proximity between product approval and publications, the relative time was assessed between the reported regulatory approval date for the product on the EEA/GB market and the date of publication (Fig. 4C-D). This showed that, for products with accompanying publications, approximately 53% (8/15) were associated with a first publication on, or prior to, the date of approval, while 47% (7/15) received approval prior to their first scientific publication (Fig. 4C-D).

**Fig. 4.**
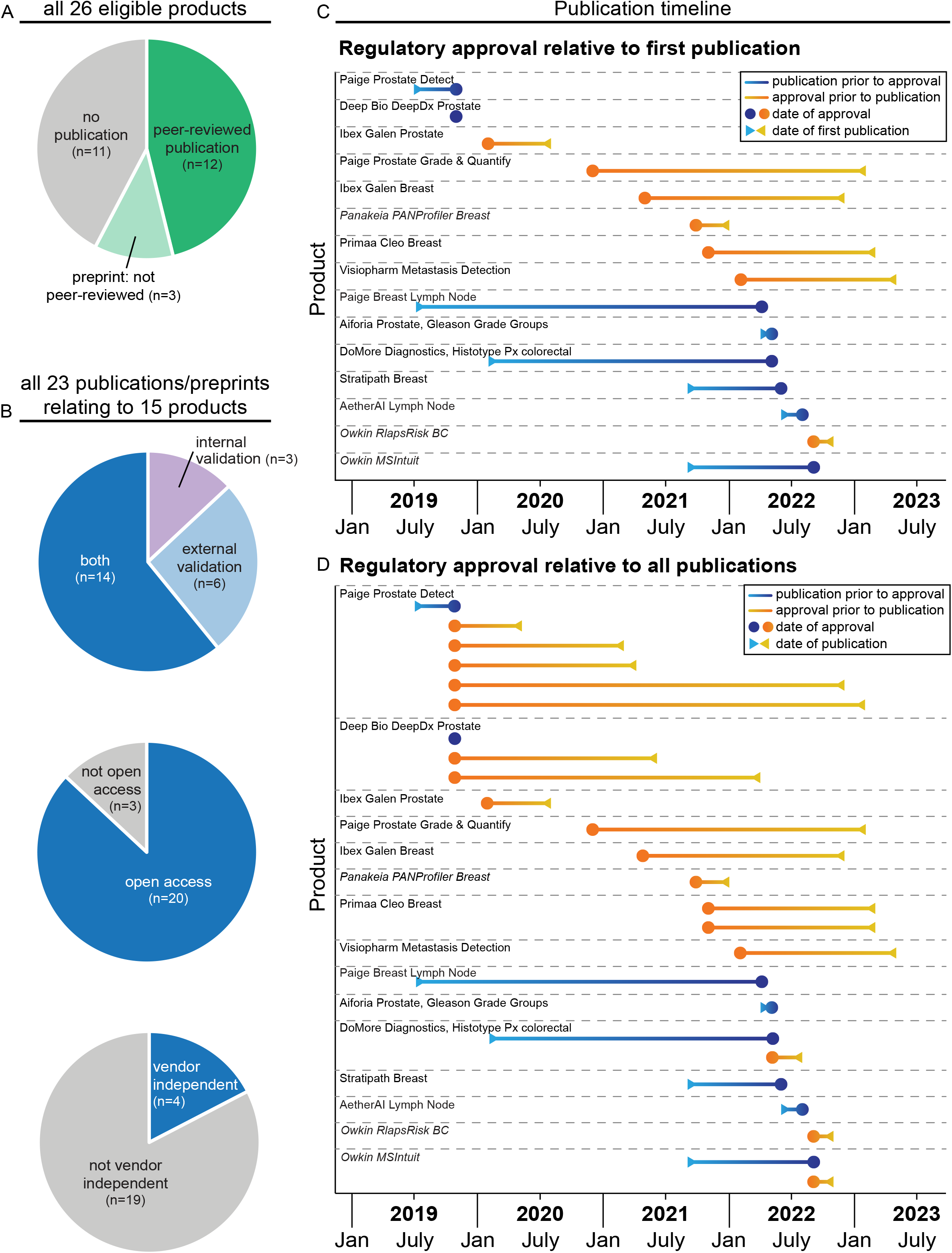
Scientific publications associated with eligible products. (**A**) Pie charts showing the proportion of products with accompanying scientific publications, (**B**) the type of study described, the accessibility of the publication, and the independence from vendors. (**C**) Graphs depicting the date between a product receiving regulatory approval and release of the first associated scientific publication, and (**D**) all scientific publications. Products with preprints are indicated in italics. Information was extracted from 23 identified publications, two of which described the validation of two separate products. Therefore 25 lines are displayed on the graph to indicate the publications related to the different products.

The design of algorithm validation studies was assessed by extracting information relating to certain study features from each available publication. This included the number of images used in training/testing datasets, the number of data sources, the scanning platforms used, and the performance metrics reported (Table S2). Two publications described studies on two different products, therefore, for analysis purposes, these were treated as separate ‘studies’ resulting in 18 studies describing internal validation and 22 studies describing external validation. For internal validation studies, which comprised 18 studies on 13 products, algorithms were reportedly trained on ∼500-10,000 WSIs images (∼150-2,500 cases/patients) and tested on ∼25-2,500 WSIs (∼25-1,500 cases/patients, Fig. 5A). Datasets for internal validation testing were typically from 1-3 sources and scanned on 1-2 scanner platforms (Fig. 5B). The datasets were obtained from a variety of countries across the 18 internal validation studies, with 17% (3/18) of these studies including test data from a UK institution (Fig. 5C-E). Thus, two products overall were associated with public evidence of internal validation on UK data.

**Fig. 5.**
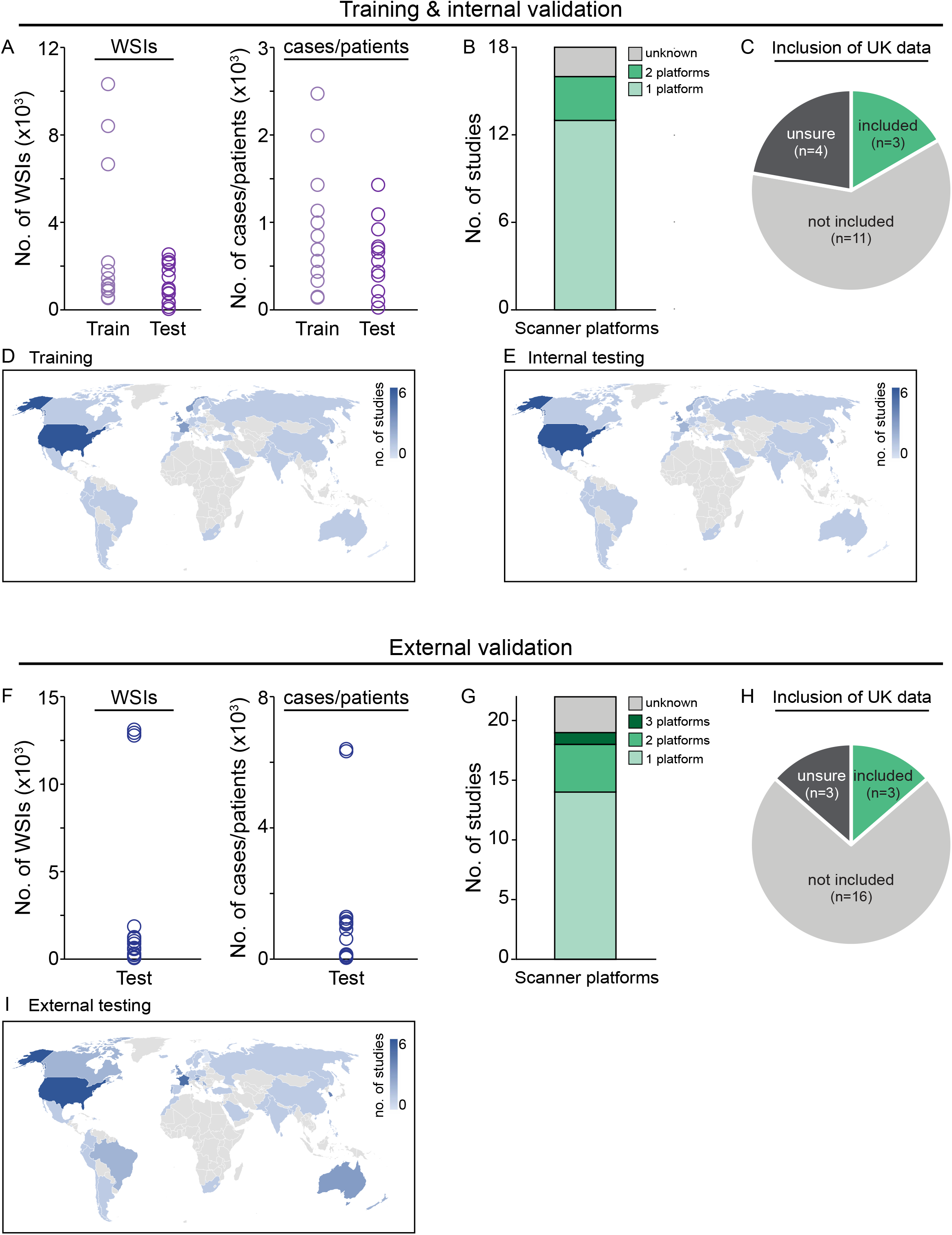
Datasets used for validation studies. (**A**) Number of whole slide images (WSIs) and cases/patients used for training and internal validation testing datasets across all studies. (**B**) Number of scanner platforms used to produce WSIs for internal validation testing data. (**C**) Pie chart showing the proportion of studies that included data from a UK-based institution in their internal validation. (**D**) World map displaying the country of origin of the data used for training and (**E**) testing for internal validation studies. (**F**) Number of WSIs and cases/patients used for external validation testing across all studies. (**G**) Number of scanner platforms used to produce WSIs for external validation testing data. (**H**) Pie chart showing the proportion of studies that included data from a UK-based institution in their external validation. (**I**) World map displaying the country of origin of the data used for external validation studies.

In the 22 external validation studies on 14 products, algorithms were tested on ∼100-13,000 WSIs (∼15-6,500 cases/patients, Fig. 5F). External validation data typically originated from 1-3 sources; however, four products were tested on data from >20 sources. Similar to internal validation, images for external validation studies were generally acquired on 1-2 scanner platforms, with the Leica Aperio AT2 being the most prominent scanner type (Fig. 5G). Datasets for external validation were obtained from a similar range of countries as for internal validation, with 14% (3/22) of the studies including data from a UK institution (Fig. 5H-I). Thus, two products overall were associated with public evidence of external validation on UK data. These data highlight the variability across validation studies used to assess the performance of AI-based products for digital pathology.

## Discussion

### Transparency surrounding AI-based medical devices for digital pathology

The rapid emergence of novel AI-based tools to support the digital pathology workflow has coincided with an increased awareness of the need for transparency, openness, and increased scrutiny around AI applications (e.g., ^31–34^). This includes a need to address not only transparency in terms of algorithmic performance, and the potential for performance drift, but also in terms of the bias present in training data. ^5,35,36^. To support decision making and public trust around the introduction of novel medical technology, information about new products needs to be both transparent and accessible to the range of stakeholders that will be impacted^19^. This is corroborated by an NHS Transformation Directorate public survey, used to inform the UK’s National Strategy for AI in Health and Social Care, which highlighted that the public want easy access to information about products compliance with medical device regulation^37^. Our analysis of AI-based products approved for the EEA/GB market identified 26 products for H&E images, 24 of which had been approved as General IVDs under the IVDD legislation, which does not require review by a notified body. Furthermore, not all manufacturers or devices could be located on public databases, and less than half of the products were associated with peer-reviewed publications describing their clinical validation. However, usefully, the majority of those publications were open access. Thus, information on medical device regulatory approval and clinical validation was not consistently available, and there was no up-to-date independent database where essential information could be verified.

The benefit of accessible information on medical devices was highlighted in one recent study in the USA, which noted that ∼19% of AI-based medical devices in radiology portrayed discrepancies between their FDA-approved indications for use and their marketing material^38^. This analysis was enabled by the publicly accessible FDA approval summaries. However, in the EEA/GB, this independent verification of vendor information would be significantly more challenging, due to the limited information on devices approved via the EEA/GB regulatory process, the lack of a comprehensive public database, and the opacity of evidence assessed by notified bodies^19,39^. This emphasises the value of sources of independently verifiable information, and the need for initiatives that support openness and transparency around novel AI medical devices.

### Public registries of medical devices

Recent changes in regulatory frameworks have recognised the need for improved information transparency and accessibility, for example the EU AI Act^40^, the introduction of the EUDAMED database, and the increased rigor of novel medical device assessment, e.g., through more products requiring notified body review under the new EU IVDR. There are also several independent initiatives supporting market transparency, for example the AIforRadiology.com public database of commercially available products in radiology^27^, and the construction of a similar registry by the Royal College of Radiologists (RCR)^28^. The RCR database has indicated it will also contain information on which hospital sites are using AI products, which will enable an understanding of the extent of use of those products and promote sharing of first-hand experience^28^.

To promote this same culture of accessibility/openness in pathology, we have constructed a register of AI products for digital pathology: https://resources.npic.uk/AI/ProductRegister. The register includes information relating to a product’s primary function, disease area, date and type of regulatory approval, and peer-reviewed clinical validation studies where available. We intend to maintain product-related information on a regular basis and expand the content and scope of the register to cover all AI-based products for image analysis in digital pathology. It is hoped this will provide a useful resource for pathologists, commissioners, and other stakeholders to easily locate essential information on products with the potential for clinical deployment.

### Quality of clinical validation studies

A major unifying challenge across AI-based medical devices is assuring high levels of accuracy and generalisability. In a typical development pipeline, new AI models undergo external validation, in which their performance is assessed on a distinct, unseen dataset^41^. External validation is critical to evaluate accuracy and generalisability, but it relies on suitably constructed test datasets, which capture the full diversity of real-world data. This is a particular challenge in digital pathology, given the variation that the slide preparation and WSI creation process can introduce into the final visualised image^5–7,42^. Importantly, this variation can impact algorithmic performance. For example, differences in scanner platforms^43^, image resolution^44^, and image quality^45^ have all been associated with differences in performance on image analysis tasks. Therefore, products which have been tested on datasets with little diversity may hold limited value in real-world clinical practice, potentially resulting in inconsistent performance across sites, confining their use to a particular scanner platform, and/or requiring site-specific re-training. Thus, well-constructed external test datasets are essential to ensure that products are only deployed into operational environments using infrastructure on which they have been validated^7^.

Our analysis showed that, of 26 regulatory approved AI products, only 42% had a peer-reviewed publication describing external validation. This is consistent with the radiology landscape, where one study identified that only 36% of commercially available AI products had peer-reviewed evidence of efficacy^26^. In our analysis of published studies, the size of test datasets for external validation was highly variable, and, where details of scanner platforms were included, we found that about two thirds of studies acquired data on just one scanner platform. Furthermore, of the 15 products associated with publications, approximately half were published after a product had received regulatory approval. Variable standards for evaluation of AI-based medical devices have also been described in other fields, including an analysis of all summaries for FDA-approved medical AI devices, which showed that only 28% reported details of a multi-site evaluation, and only 3% used prospective data^29^. However, encouragingly, an analysis of the radiology AI market showed that, while only 30% and 25% of individual studies used multicentre and multi-scanner data, respectively^26^, when aggregating by product, the majority were validated on multi-site and multi-scanner data^26^.

The variability and opacity of clinical evaluation for market approval necessitates the development of specific standards for algorithm validation studies. Despite general guidance indicating that devices must show high levels of accuracy, and be tested on representative datasets (e.g.,^1–4^), further work is needed to establish best practices for compiling test datasets and evaluating AI-based medical devices for digital pathology^5,29,46,47^. An additional feature that emerged from our analysis was that, of the 16 products that were UKCA-marked and/or CE-marked with an MHRA-registered manufacturer, only two had publications reporting the use of UK data for internal or external validation. The lack of product testing on UK data has been highlighted by NICE, in their Medtech Innovation Briefing for an AI product in pathology^25^, and mammography^48^, as well as their Diagnostics Guidance on AI software for lung nodule detection on CT^49^. This suggests exercising caution in these circumstances, and highlights the need for further testing on UK-based data, which is currently underway alongside deployment for several of the more advanced AI products for digital pathology (e.g., ^50,51^).

### Limitations

Our focus in this study was to identify and analyse evidence on AI-based products for digital pathology that was publicly accessible. Therefore, it is possible that some information was missed if it was not publicly available or was inaccessible. In many cases, we found that the product information provided on company public websites was limited, hard to locate, or ambiguously presented. In addition, websites sometimes required entry of personal information (e.g., name, email address) to request further details on a product. Therefore, there may be additional clinical evidence that has been gathered in support of these products, but it could not be included here if it was not available to the public. Furthermore, in this study, we limited our inclusion criteria to products which used only H&E images as their input, as this is the most common stain used in histopathology. However, we intend to broaden the scope of the public register to include AI products that make predictions from images with other stains or immunohistochemistry.

## Conclusions

Pathology is the cornerstone of cancer diagnosis and therefore changes to this workflow could have a significant impact on patient outcomes. Although AI-based image analysis products have the potential to revolutionise the diagnostic workflow, the cost and resources involved in clinical deployment is high, and therefore products with a strong evidence base would be more likely to result in successful clinical adoption. In our analysis, we found that public evidence on regulatory approved AI-based products for digital pathology was often limited and difficult to access, while independently reviewed clinical evidence was available for less than half of the products. Providing clear, accessible information on novel medical devices is essential in promoting public trust and facilitating informed judgements by healthcare professionals and commissioners. Product vendors may be understandably hesitant to share information on their products at an early stage, due to concerns around compromising their market position. However, increased openness and transparency of evidence relating to marketable medical devices will benefit all stakeholders, including vendors, as all parties can quickly and easily locate available products and verify information. Public registries can support this endeavour, by providing up to date information as the market evolves and offering the capacity for long-term monitoring as products are deployed across the NHS.

## Data Availability

All data produced in the present work are contained in the manuscript.

## Author Contributions

G.A.M. and D.T. designed the study. G.A.M. conducted the searches and G.A.M and C.M. performed data extraction. G.A.M. analysed the data and wrote the manuscript. All authors reviewed and edited the manuscript.

