## Supplementary Table S1 and S2 for "Public evidence on AI products for digital pathology"

### Supplementary Materials

Table S1……………………………………………………………………………………………………………………… 2

Table S2……………………………………………………………………………………………………………………… 5

Glossary & Abbreviations……………………………………………………………………………………………...… 11

Supplementary References…………………………………………………………………………………………..…… 12

**Table S1**: Table summarising the information gathered for each product that met criteria for inclusion.

| Product name | Company details | | | Product details | | | | Regulatory information | | | | | | Publications | |
|---|---|---|---|---|---|---|---|---|---|---|---|---|---|---|---|
| | Company name | Location | Size | Tissue type | Pathology Subspecialty | Primary Purpose | Key features/ suggested use | Regulatory approval type and class | Approx date of approval | MHRA PARD | EUDAMED actor | EUDAMED device | FDA database | Publications: internal validation | Publications: external validation |
| **AetherAI LN, Lymph Node Metastasis AI Detection** | AetherAI | Taipei, Taiwan | 51-200 employees | lymph nodes | Gastrointestinal pathology | Detection with quantification | Detects metastases in lymph nodes from patients with gastric cancer and classifies/quantifies positive and negative lymph nodes. | CE-IVD General (IVDD) | Aug 2022 (estimated) | N | N | N | N | 1 | 1 |
| **Aiforia Clinical AI Model for Prostate Cancer, Gleason Grade Group** | Aiforia | Helsinki, Finland | 51-200 employees | prostate | Uropathology | Detection with grading | Detects tumour areas and predicts Gleason score and grade group. | CE-IVD General (IVDD) | May 2022 | N | Y | Y | N | 1 | 0 |
| **DeepDx Prostate** | Deep Bio | Seoul, South Korea | 11-50 employees | prostate | Uropathology | Detection with grading & quantification | Detects tumour areas and predicts Gleason score and grade group. Quantifies proportion of each Gleason pattern out of total tumour area and tumour-to-tissue ratios. | CE-IVD General (IVDD); MFDS (Korea) Class III IVD | Nov 2019 | N | N | N | N | 2 | 2 |
| **DeepPath LYDIA** | DeepMed IO | Manchester, UK | 2-10 employees | lymph nodes | Multiple | Detection | Detects metastases in lymph nodes from patients with melanoma, breast, colon, and lung cancer. | CE-IVD General (IVDD) | May 2022 | N | N | N | N | 0 | 0 |
| **Histotype Px colorectal** | DoMore Diagnostics | Oslo, Norway | 2-10 employees | colorectal | Gastrointestinal pathology | Prognosis/ treatment selection | Predicts risk profile and cancer-specific survival of stage II & III colorectal cancer patients. | CE-IVD General (IVDD) | May 2022 | N | Y | N | N | 2 | 2 |
| **Galen Breast** | Ibex Medical Analytics | Tel Aviv, Israel | 51-200 employees | breast | Breast pathology | Detection with grading & quantification | Detects tumour area and predicts Gleason grade group, and cancer subtype. Quantifies tumour measurements and detects other clinically significant features, e.g., tumour-infiltrating lymphocytes (TILs), lymphovascular invasion, and microcalcifications. | CE-IVD General (IVDD) | May 2021 | Y | Y | N | N | 1 | 1 |
| **Galen Prostate** | Ibex Medical Analytics | Tel Aviv, Israel | 51-200 employees | prostate | Uropathology | Detection with grading & quantification | Detects tumour area and predicts Gleason grade group. Quantifies tumour measurements and detects other clinically relevant features, e.g., perineural invasion. | CE-IVD General (IVDD) – Galen Prostate; CE-IVD Class C (IVDR) – Galen First Read for Prostate | Feb 2020 (IVDD) Feb 2023 (IVDR) | Y | Y | N | N | 1 | 1 |
| **Galen Gastric** | Ibex Medical Analytics | Tel Aviv, Israel | 51-200 employees | stomach | Gastrointestinal pathology | Detection with grading & quantification | Detects carcinoma, high- and low-grade dysplasia, Helicobacter pylori (H. pylori), neuroendocrine lesions, adenoma, and other features. | CE-IVD General (IVDD) | June 2022 | Y | Y | N | N | 0 | 0 |
| **HALO Prostate AI** | Indica Labs | New Mexico, USA | 51-200 employees | prostate | Uropathology | Detection with grading & quantification | Detects tumour area and predicts Gleason grade group. Quantifies tumour measurements and detects other clinically relevant features, e.g., high-grade Prostatic Intraepithelial neoplasia (PIN) and intraductal carcinoma. | CE-IVD General (IVDD) | May 2022 | Y | Y | N | N | 0 | 0 |
| **INIFY Prostate** | INIFY Laboratories | Stockholm, Sweden | 11-50 employees | prostate | Uropathology | Detection | Detects tumour area and quantifies tumour volume. | CE-IVD General (IVDD) | June 2020 | N | N | N | N | 0 | 0 |
| **RlapsRisk BC** | Owkin | Paris, France | 201-500 employees | breast | Breast pathology | Prognosis/ treatment selection | Predicts probability of relapse at 5yrs for patients with HER2-/ER+ primary invasive breast cancer. | CE-IVD General (IVDD)[a] | Sept 2022 | Y | Y | N | N | 1 | 1 |
| **MSIntuit CRC** | Owkin | Paris, France | 201-500 employees | colorectal | Gastrointestinal pathology | Biomarker Prediction | Detects microsatellite instability (MSI) in colorectal cancer in order to rule out microsatellite stable (MSS) phenotypes and aid clinical management decisions. | CE-IVD General (IVDD) | Sept 2022 | Y | Y | N | N | 2 | 2 |

[a] Website indicates '*The device is currently under development, and not for clinical use*'.

| Product name | Company details | | | Product details | | | | Regulatory information | | | | | | Publications | |
|---|---|---|---|---|---|---|---|---|---|---|---|---|---|---|---|
| | Company name | Location | Size | Tissue type | Pathology Subspecialty | Primary Purpose | Key features/ suggested use | Regulatory approval type and class | Approx date of approval | MHRA PARD | EUDAMED actor | EUDAMED device | FDA database | Publications: internal validation | Publications: external validation |
| **Paige Breast Detect & Neoplasm** | Paige.AI | NY, USA | 51-200 employees | breast | Breast pathology | Detection with grading | Detects and classifies malignant neoplasms and predicts neoplasm subtype. | CE-IVD General (IVDD) & UKCA[b] | Dec 2020 (IVDD) - Detect. June 2023 (IVDD & UKCA) - Breast Suite[c] | Y | Y | N | N | 0 | 0 |
| **Paige Breast Lymph Node** | Paige.AI | NY, USA | 51-200 employees | breast | Breast pathology | Detection | Detects metastases and isolated tumour cells (ITCs) in lymph nodes from patients with breast cancer. | CE-IVD General (IVDD) & UKCA[b] | April 2022 | Y | Y | N | N | 1 | 1 |
| **Paige Breast Mitosis** | Paige.AI | NY, USA | 51-200 employees | breast | Breast pathology | Detection with quantification | Detects and quantifies mitoses within invasive cancer regions. | CE-IVD General (IVDD) & UKCA[b] | June 2023 (estimated) | Y | Y | N | N | 0 | 0 |
| **HER2Complete BETA** | Paige.AI | NY, USA | 51-200 employees | breast | Breast pathology | Biomarker Prediction | Predicts HER2 expression in tumour regions. | CE-IVD General (IVDD) & UKCA[b,d] | June 2022 | Y | Y | N | N | 0 | 0 |
| **Paige Prostate Detect** | Paige.AI | NY, USA | 51-200 employees | prostate | Uropathology | Detection | Detects foci suspicious for cancer and provides slide-level binary classification. | CE-IVD General IVD (IVDD) & UKCA[e]; FDA class II IVD[f] | Nov 2019 (IVDD) Sept 2021 (FDA) | Y | Y | N | N | 2 | 5 |
| **Paige Prostate Grade & Quantify** | Paige.AI | NY, USA | 51-200 employees | prostate | Uropathology | Detection with grading & quantification | Quantifies tumour measurements and predicts Gleason score and Gleason pattern for suspicious areas. | CE-IVD General (IVDD) & UKCA[g] | Dec 2020 | Y | Y | N | Y | 0 | 1 |
| **Paige Prostate PNI** | Paige.AI | NY, USA | 51-200 employees | prostate | Uropathology | Detection | Detects suspicious foci around nerve fibres to assist in diagnosis of perineural invasion (PNI). | CE-IVD General (IVDD) & UKCA[g] | Feb 2022 (estimated) | Y | Y | N | N | 0 | 0 |
| **Paige Prostate Biomarker Suite** | Paige.AI | NY, USA | 51-200 employees | prostate | Uropathology | Biomarker Prediction | Detects areas with increased likelihood of Androgen Receptor (AR) amplification, and TP53, RB1 and PTEN mutations. | CE-IVD General (IVDD) & UKCA[g] | May 2022 | Y | Y | N | N | 0 | 0 |
| **PANProfiler Breast** | Panakeia Technologies | Cambridge, UK | 11-50 employees | breast | Breast pathology | Biomarker Prediction | Predicts the status of biomarkers ER, PR, and HER2. | CE-IVD General (IVDD) & UKCA | Oct 2021 | Y | Y | N | N | 1 | 1 |
| **Cleo Breast Cancer** | Primaa | Paris, France | 11-50 employees | breast, lymph nodes | Breast pathology | Detection with grading & quantification | Detects tumour area in breast tissue, detects metastases in lymph nodes, performs mitosis counting, predicts Nottingham grade, and identifies clinically relevant features e.g., calcification. | CE-IVD General (IVDD) | Nov 2021 | Y | Y | N | N | 2 | 2 |

[b] Approved on Leica Aperio AT2 and GT450 scanners.

[c] Paige Breast Suite consists of: Paige Breast Detect & Neoplasm, Paige Breast Mitosis, Paige Breast Lymph Node and HER2Complete.

[d] Released for product evaluation and not currently available for purchase.

[e] Approved on Leica Aperio AT2 and Philips Ultrafast scanners.

[f] Approved on Philips Ultra Fast scanner with Paige FullFocus WSI viewing software.

[g] Approved on Leica Aperio AT2 and Philips Ultra Fast Scanners.

| Product name | Company details | | | Product details | | | | Regulatory information | | | | | | Publications | |
|---|---|---|---|---|---|---|---|---|---|---|---|---|---|---|---|
| | Company name | Location | Size | Tissue type | Pathology Subspecialty | Primary Purpose | Key features/ suggested use | Regulatory approval type and class | Approx date of approval | MHRA PARD | EUDAMED actor | EUDAMED device | FDA database | Publications: internal validation | Publications: external validation |
| **Stratipath Breast** | Stratipath | Stockholm, Sweden | 11-50 employees | breast | Breast pathology | Prognosis/ treatment selection | Detects tumour area and predicts risk of recurrence for intermediate Nottingham Grade 2 tumours. | CE-IVD General (IVDD) | June 2022 | N | Y | N | N | 1 | 1 |
| **Metastasis Detection, AI** | Visiopharm | Hoersholm, Denmark | 51-200 employees | lymph nodes | Multiple | Detection with quantification | Detects metastases in lymph nodes, and measures and orders metastases based on relevant features. | CE-IVD Class C (IVDR) | Feb 2020 (IVDD) Feb 2022 (IVDR) | Y | Y | N | N | 0 | 1 |
| **Zeno Pathology: Her2** | WSK Medical | Amsterdam, Netherlands | 2-10 employees | breast | Breast pathology | Biomarker Prediction | Predicts HER2 score (negative, equivocal, positive) | CE-IVD General (IVDD) | uncertain | N | Y | N | N | 0 | 0 |
| **Zeno Pathology: lymph node metastasis** | WSK Medical | Amsterdam, Netherlands | 2-10 employees | lymph nodes | Other | Detection | Detects metastases in lymph nodes | CE-IVD General (IVDD) | uncertain | N | Y | N | N | 0 | 0 |

**Table S2**: Table summarising the information extracted from all available scientific publications related to each product.

| General Details | | | | | | | | Training and Internal Validation | | | | | | | External Validation | | | | |
|---|---|---|---|---|---|---|---|---|---|---|---|---|---|---|---|---|---|---|---|
| **Company name** | **Product name** | **Paper title** | **DOI** | **Date** | **Vendor independence** | **Open access** | **Internal/ external validation** | **Training set:** no. WSIs, cases, patients | **Test set:** no. WSIs, cases, patients | **Training set:** no. data sources, countries | **Test set:** no. data sources, countries | **UK data** | **Test set:** scanner platforms | **Performance** | **Test set:** no. WSIs, cases, patients | **Test set:** no. data sources, countries | **UK data** | **Test set:** scanner platforms | **Performance** |
| **AetherAI** | **AetherAI LN** | Deep neural network trained on gigapixel images improves lymph node metastasis detection[1] | https://doi.org/10.1038/s41467-022-30746-1 | Jun-22 | No | Yes | Both | 1,093 WSIs<br>(Training: 983 WSI; validation & tuning: 110 WSIs) | Main test set: 201 WSIs;<br>Clinical test set: 80 WSIs | 1,1 (Taiwan) | 1,1 (Taiwan) | No | 1 (Hamamatsu NanoZoomer S360) | WSI-level performance, main test set:<br>Classification of positive/negative lymph nodes (LNs): AUC=0.9936.<br>LN image-level performance: Classification of LNs: sens=0.8915; spec=0.9861, PPV=0.9564, NPV=0.9637; MCC=0.8986.<br>Clinical test set: Pathologist median WSI review time (without vs with AI): 161.2s vs 110.5s. | 327 WSIs | 1,1 (Taiwan) | No | 1 (Hamamatsu NanoZoomer S360) | WSI-level performance:<br>Classification of positive/negative LNs: AUC=0.9829 |
| **Aiforia** | **Aiforia Clinical AI Model for Prostate Cancer, Gleason Grade Group** | AI Model for Prostate Biopsies Predicts Cancer Survival[2] | https://doi.org/10.3390/diagnostics12051031 | Apr-22 | No | Yes | Internal | 516 WSIs, 331 patients | 2088 WSIs, 391 patients | 1,1 (Finland) | 1,1 (Finland) | No | 1 (3DHistech Panoramic 250 Flash III) | Biopsy level performance:<br>Classification (benign vs malignant): AUC=0.997; acc=0.98, sens=0.98, spec=0.98, PPV=0.96, NPV=0.99. Concordance with pathologists: weighted κ=0.96;<br>Patient level performance:<br>[h]Grade Group classification: acc=0.67, sens=0.11-0.98, spec=0.87-0.98, PPV=0.09-0.96, NPV=0.85-0.99. Concordance with pathologists: weighted κ=0.766. | NA | NA | NA | NA | NA |
| **Deep Bio** | **DeepDx Prostate** | Automated Gleason Scoring and Tumor Quantification in Prostate Core Needle Biopsy Images Using Deep Neural Networks and Its Comparison with Pathologist-Based Assessment[3] | https://doi.org/10.3390/cancers11121860 | Nov-19 | No | Yes | Internal | 1,133 WSIs, 1,133 cases | 700 WSIs, 700 cases | 2,1 (South Korea) | 2,1 (South Korea) | No | 1 (Leica Aperio AT2 scanner) | WSI/case-level performance:<br>Grade Group classification: concordance with pathologists: κ=0.615, quadratic weighted κ=0.907.<br>Tumour length measurements: concordance with pathologists: r=0.97. | NA | NA | NA | NA | NA |
| **Deep Bio** | **DeepDx Prostate** | Yet Another Automated Gleason Grading System (YAAGGS) by weakly supervised deep learning[4] | https://doi.org/10.1038%2Fs41746-021-00469-6 | Jun-21 | No | Yes | Both | 6664 WSIs, 689 cases/patients<br>(Training: 5,716 WSIs; tuning: 948 WSIs) | 936 WSIs, 99 cases/patients | 2,1 (South Korea) | 2,1 (South Korea) | No | 1 (Leica Aperio AT2 scanner) | WSI-level performance:<br>Cancer detection: AUC=0.983, AUPRC=0.984, acc=0.947, sens=0.936, spec=0.960.<br>Grade group classification: acc=0.775; concordance with pathologists: κ=0.650, quadratic weighted κ=0.897. | 244 WSIs (Tissue Microarrays) | 1,1 (Canada) | No | 1 (Leica SCN400 scanner) | WSI-level performance:<br>Cancer detection: AUC=0.943, AUPRC=0.985. Grade group classification: acc=0.545, concordance with pathologists: κ=0.389, quadratic weighted κ=0.634. |
| **Deep Bio** | **DeepDx Prostate** | Artificial intelligence system shows performance at the level of uropathologists for the detection and grading of prostate cancer in core needle biopsy: an independent external validation study[5] | https://doi.org/10.1038/s41379-022-01077-9 | Apr-22 | Yes | Yes | External | NA | NA | NA | NA | NA | NA | NA | 593 WSIs/cases | 1,1 (South Korea) | No | 1 (Leica Aperio AT2) | WSI-level performance:<br>Cancer detection: acc=0.9831, sens=0.9870, spec=0.9692, PPV=0.9913, NPV=0.9545. Grade Group classification: concordance with pathologists: κ=0.713, quadratic weighted κ=0.922; Grade Score classification: concordance with pathologists: κ=0.654, quadratic weighted κ=0.904, Spearman's rho=0.938;<br>Pathologist performance (without vs with AI): Cancer detection: acc=0.9612 vs 0.9831, sens=0.9957 vs 0.9849, spec=0.8385 vs 0.9767, PPV=0.9564 vs 0.9935, NPV=0.9820 vs 0.9474. Grade Group concordance: κ=0.621 vs 0.741, quadratic weighted κ=0.876 vs 0.925. Average examination time: 55.7s vs 36.8s/case. |

[h] Classification performance across all classes (i.e., benign and Grade Groups 1-5) is presented, therefore a range of values is displayed.

| General Details | | | | | | | | Training and Internal Validation | | | | | | | External Validation | | | | |
|---|---|---|---|---|---|---|---|---|---|---|---|---|---|---|---|---|---|---|---|
| **Company name** | **Product name** | **Paper title** | **DOI** | **Date** | **Vendor independence** | **Open access** | **Internal/ external validation** | **Training set:** no. WSIs, cases, patients | **Test set:** no. WSIs, cases, patients | **Training set:** no. data sources, countries | **Test set:** no. data sources, countries | **UK data** | **Test set:** scanner platforms | **Performance** | **Test set:** no. WSIs, cases, patients | **Test set:** no. data sources, countries | **UK data** | **Test set:** scanner platforms | **Performance** |
| **DoMore Diagnostics** | [i]**Histotype Px colorectal** | Deep learning for prediction of colorectal cancer outcome: a discovery and validation study[6] | https://doi.org/10.1016/S0140-6736(19)32998-8 | Feb-20 | [j]Yes | No | Both | 2473 patients (828 training, 1645 tuning) | 920 patients | [k]154, 2 (Norway, UK) | 1,1 (UK) | Yes: train & test | 2 (Leica Aperio AT2, Hamamatsu NanoZoomer XR) | All patients, cancer-specific survival prognosis:<br>Leica: uncertain vs good prognosis HR=2.54; poor vs good prognosis HR=4.83.<br>Hamamatsu: uncertain vs good prognosis HR=2.03, poor vs good prognosis HR=3.98.<br><br>Harrell's concordance index (C-index) between model prediction and cancer-specific survival = 0.695 (Leica), 0.692 (Hamamatsu).<br><br>Patients with distinct outcomes (i.e., good or poor): AUC=0.718 (Leica), AUC=0.715 (Hamamatsu). | 1122 patients | [l]170, 7 (Australia, Austria, Czech Republic, New Zealand, Serbia, Slovenia, UK) | Yes | 2 (Leica Aperio AT2, Hamamatsu NanoZoomer XR) | Stage II & III patients, cancer-specific survival prognosis:<br>Leica: uncertain vs good prognosis HR=1.89; poor vs good HR=3.84. Multivariable analysis, after adjusting for established prognostic markers, uncertain vs good HR=1.56, poor vs good HR=3.04.<br>Hamamatsu: uncertain vs good HR=2.42, poor vs good HR=3.39. Multivariable analysis, uncertain vs good HR=1.80, poor vs good HR=2.46.<br>Harrell's concordance index between model prediction and cancer-specific survival = 0.674 (Leica), 0.674 (Hamamatsu).<br>Patients with distinct outcomes (i.e., good or poor): AUC=0.713 (Leica), 0.724 (Hamamatsu)<br>Leica: Prediction of 3yr cancer-specific survival for good vs poor & uncertain prognosis: acc (proportion of correctly classified patients)=0.76, sens=0.52, spec=0.78, PPV=0.19, NPV=0.94.<br>Prediction of 3yr cancer-specific survival for good & uncertain vs poor prognosis: acc=0.67, sens=0.69, spec=0.66, PPV=0.17, NPV=0.96. |
| **DoMore Diagnostics** | **Histotype Px colorectal** | A clinical decision support system optimising adjuvant chemotherapy for colorectal cancers by integrating deep learning and pathological staging markers: a development & validation study[7] | https://doi.org/10.1016/S1470-2045(22)00391-6 | Aug-22 | No | No | Both | [m]997 patients | [n]657 patients (subset of training data) | 2,2 (Norway, UK) | 2,2 (Norway, UK) | Yes: train & test | 1 (Leica Aperio AT2) | Cancer-specific survival prognosis:<br>Low vs intermediate risk HR=3.56, low vs high risk = 11.69.<br><br>Estimated 3 yr cancer-specific survival = 96.2% for low risk, 85.1% for intermediate risk, and 50.5% for high risk. | 1075 patients | [o]170, 7 (Australia, Austria, Czech Republic, New Zealand, Serbia, Slovenia, UK) | Yes | 1 (Leica Aperio AT2) | Cancer-specific survival prognosis:<br>Low vs intermediate risk HR=3.06, low vs high risk HR=10.71.<br><br>Estimated 3yr cancer-specific survival = 97.2% for low risk, 94.8% for intermediate risk, 77.6% for high risk. |
| **Ibex Medical Analytics** | [p]**Galen Breast** | Validation and real-world clinical application of an artificial intelligence algorithm for breast cancer detection in biopsies[9] | https://doi.org/10.1038/s41523-022-00496-w | Dec-22 | No | Yes | [q]Both | 2,153 WSIs, 1,992 patients | 2,252 WSIs, 1,090 patients | [r]9, unknown | 1,1 (Israel) | No | 1 (Philips IntelliSite Scanner) | Case-level performance:<br>Carcinoma detection (invasive vs non-invasive): AUC=0.998, sens=0.9902, spec=0.9827, PPV=0.95, NPV=0.997.<br><br>Subtype differentiation: ductal carcinoma in situ (DCIS) vs benign/other non-invasive tumours: AUC=0.999, sens=1.00, spec=0.9864, PPV=0.693, NPV=1.00; invasive ductal carcinoma (IDC) vs invasive lobular carcinoma (ILC): AUC=0.932. | External validation: 841 WSIs, 436 patients;<br><br>Second read clinical deployment: 12,031 WSIs, 5,954 cases | 2,2 (Israel, France) | No | 2 (Hamamatsu NanoZoomer S360; Philips IntelliSite Scanner) | Case-level performance:<br><br>Carcinoma detection (invasive vs non-invasive): AUC=0.990, sens=0.9551, spec=0.9357, PPV=0.892, NPV=0.974.<br>DCIS vs benign/other non-invasive tumours: AUC=0.980, sens=0.9320, spec=0.9379, PPV=0.914, NPV=0.951.<br><br>Subtype/grade differentiation:<br>rare invasive vs non-invasive: AUC=0.992, AUPRC=0.9434, sens=0.9571, spec=0.9412, PPV=0.727, NPV=0.993, $F_1$=87.5%;<br>DCIS/Atypical Ductal Hyperplasia (ADH) vs benign/other: AUC=0.949, sens=0.8741, spec=0.869, PPV=0.861, NPV=0.881;<br>IDC vs ILC: AUC=0.973, sens=0.9286, spec=0.9273, PPV=0.958, NPV=0.879;<br>DCIS high grade/intermediate grade vs low grade/ADH: AUC=0.921, sens=0.8478, spec=0.8409, PPV=0.914, NPV=0.736;<br><br>Detection of Tumour Infiltrating Lymphocytes (TILs) in invasive & DCIS: AUROC=0.965, sens=0.938, spec=0.857.<br><br>Second read clinical deployment:<br>Invasive carcinoma detection: AUC=0.990, sens=0.9809, spec=0.9624.<br>DCIS/ADH detection: AUC=0.972, sens=0.9230, spec=0.9222. |

[i] Publication notes that "*The early version of Histotype Px® Colorectal was named DoMore-V1-CRC (and misc. variations of this description) during development in the DoMore! Project. The CE-IVD certified product Histotype Px® Colorectal is an updated version*".

[j] Classed as independent of vendors as company formation occurred after publication.

[k] Internal validation included four datasets, one of which was a clinical trial involving 151 UK hospitals. Slides across the four datasets were prepared at two locations.

[l] External validation was performed on one dataset from the QUASAR-2 clinical trial that involved 170 hospitals across seven countries.

[m] Training data used to define risk stratification system. This included data used in training/testing in the previous publication.

[n] Internal validation performance results based on a subset of the 997 patients in the training data that did not receive adjuvant chemotherapy.

[o] External validation was performed on one dataset from the QUASAR-2 clinical trial that involved 170 hospitals across seven countries. This is presumed to be the same dataset as in the previous publication.

[p] Publication describes updating of the algorithm introduced in Pantanowitz *et al.*, 2020[8].

[q] External validation dataset includes data from Maccabi Healthcare Services (MHS), which was also the source of the internal validation test data and the source of training data for the original model described in Pantanowitz *et al.*, 2020[8], but from a different time period. All data for the second read clinical deployment study was from MHS.

[r] Training data collected from nine labs of unspecified location.

| General Details | | | | | | | | Training and Internal Validation | | | | | | | External Validation | | | | |
|---|---|---|---|---|---|---|---|---|---|---|---|---|---|---|---|---|---|---|---|
| **Company name** | **Product name** | **Paper title** | **DOI** | **Date** | **Vendor independ-ence** | **Open access** | **Internal/ external validation** | **Training set:** no. WSIs, cases, patients | **Test set:** no. WSIs, cases, patients | **Training set:** no. data sources, countries | **Test set**: no. data sources, countries | **UK data** | **Test set**: scanner platforms | **Performance** | **Test set:** no. WSIs, cases, patients | **Test set:** no. data sources, countries | **UK data** | **Test set:** scanner platforms | **Performance** |
| **Ibex Medical Analytics** | **Galen Prostate** | An artificial intelligence algorithm for prostate cancer diagnosis in whole slide images of core needle biopsies: a blinded clinical validation and deployment study[8] | https://doi.org/10.1016/s2589-7500(20)30159-x | Aug-20 | No | Yes | Both | 549 WSIs, 138 cases | 2,501 WSIs, 210 cases | 1,1 (Israel) | 1,1 (Israel) | No | 1 (Philips IntelliSite Scanner) | WSI-level performance:<br>Cancer detection: AUC=0.997, sens=0.9959, spec=0.9014, PPV=0.717, NPV=0.999 | External validation: 1,627 WSIs, 100 cases (355 parts); Algorithm calibration: 32 cases.<br>[s]Second read clinical deployment: 11,429 WSIs, 941 cases | 1,1 (USA) | No | 1 (Leica Aperio AT2) | Part-level classification:<br>Cancer detection: AUC=0.991, sens=0.9846, spec=9733, PPV=0.955, NPV=0.991; Grade differentiation: low grade (Gleason score 6 or atypical small acinar proliferation (ASAP)) vs high grade (Gleason score 7-10): AUC=0.941, sens=0.859, spec=0.9041, PPV=0.905, NPV=0.857; ASAP or Gleason Pattern (GP) 3 or 4 vs GP5: AUC=0.971; spec=90.84%, sens=0.85; PPV=0.586, NPV=0.975;<br>Perineural Invasion (PNI) detection: AUC=0.957, sens=0.8696, spec=0.9074, PPV=0.80, NPV=0.942. Cancer % estimation, concordance with pathologist: r=0.882; mean bias=-4.14% |
| **[t]Owkin** | **RlapsRisk BC** | Deep Learning Allows Assessment of Risk of Metastatic Relapse from Invasive Breast Cancer Histological Slides[10] | https://doi.org/10.1101/2022.11.28.518158 | Nov-22 | No | Yes | Both | 1429 patients | cross-validation using training dataset | 1,1, (France) | 1,1, (France) | No | 1 (Olympus VS120 scanner) | Combined model:<br>Contribution of RlapsRisk score to prognosis (metastasis-free survival; MFS), HR=1.27; model C-index=0.80. | 889 WSIs/patients | 27, 1 (France) | No | unknown | Combined model: Contribution of RlapsRisk score to prognosis (MFS), HR=1.38; model C-index=0.80.<br>Classification of patients (prognostic discrimination): RlapsRisk classifier alone: % of patients with MFS event in Low Risk group=1.42%; in High Risk group=7.95%. Combined Model Classifier: % of patients with MFS event in Low Risk group=1.22%; in High Risk group=11.26%.<br>RlapsRisk Classifier: Low Risk vs High Risk HR=4.36. Combined Model Classifier: HR=6.99.<br>RlapsRisk Classifier: cumulative sens=0.77, dynamic spec=0.67; Combined classifier: cumulative sens=0.76, dynamic spec=0.76 |
| **[t]Owkin** | **MSIntuit** | Self-supervised learning improves dMMR/MSI detection from histology slides across multiple cancers[11] | https://doi.org/10.48550/arXiv.2109.05819 | Sept-21 | No | Yes | Both | 555 patients | cross validation using training dataset | 36,1 (USA) | 36,1 (USA) | No | unknown | AUC=0.88 | 47 patients | 3,1 (South Korea) | No | 1 (Leica Aperio AT2)[u] | AUC=0.97 |
| **[t]Owkin** | **MSIntuit** | Blind validation of MSIntuit, an AI-based pre-screening tool for MSI detection from histology slides of colorectal cancer[12] | https://doi.org/10.1101/2022.11.17.22282460 | Nov-22 | No | Yes | Both | 859 WSIs, 434 patients | cross validation using training dataset | [v]24, 1 (USA) | 24,1 (USA) | No | 1 (Aperio) | Microsatellite Instability (MSI) status prediction:<br>AUC=0.93 | [w]PAIP dataset: 47 patients.<br>MPATH dataset (replicated across two scanners): Philips IntelliSite - 588 WSIs/patients; Roche DP200 - 571 WSIs/patients. | 4,2 (South Korea, France) | No | 3 (Aperio AT2, Philips-IntelliSite Ultra Fast Scanner, Roche Ventana DP200) | MSI status prediction:<br>PAIP: AUC=0.97; MPATH: Philips-UFS: AUC=0.86; sens=0.95, spec=0.47, NPV=0.9; MPATH: Roche DP200: AUC=0.88; sens=0.97, spec=0.46, NPV=0.99; |

[s] Standalone performance not reported for second read clinical deployment dataset.
[t] Preprint, not peer reviewed.
[u] Scanner model assumed from same dataset used in Saillard *et al.*, 2022[12].
[v] TCGA dataset used for algorithm training, which includes data from 24 labs.
[w] External validation included PAIP and MPATH datasets: PAIP dataset described as 'independent development cohort' with data originating from three centres in South Korea and scanned on an Aperio AT2 scanner. MPATH dataset was collected from one centre in France and replicated across two scanners (Philips IntelliSite Ultra-Fast Scanner and Roche Ventana DP200). The numbers shown include 30 WSIs used for model calibration on each scanner.

| General Details | | | | | | | | Training and Internal Validation | | | | | | | External Validation | | | | |
|---|---|---|---|---|---|---|---|---|---|---|---|---|---|---|---|---|---|---|---|
| **Company name** | **Product name** | **Paper title** | **DOI** | **Date** | **Vendor independence** | **Open access** | **Internal/ external validation** | **Training set:** no. WSIs, cases, patients | **Test set:** no. WSIs, cases, patients | **Training set:** no. data sources, countries | **Test set**: no. data sources, countries | **UK data** | **Test set**: scanner platforms | **Performance** | **Test set:** no. WSIs, cases, patients | **Test set:** no. data sources, countries | **UK data** | **Test set:** scanner platforms | **Performance** |
| **Paige.AI** | **Paige Breast Lymph Node** | Clinical-grade computational pathology using weakly supervised deep learning on whole slide images[13] | https://doi.org/10.1038/s41591-019-0508-1 | Jul-19 | No | Yes | Both | 8,421 WSIs (estimated) | 1,473 WSIs | [x]800+, 45 | [x]800+, 45 | Yes: train & test | 1 (Leica Aperio AT2; Philips IntelliSite Ultra Fast) | Metastasis detection: AUC=0.965 | [y]129 WSIs | 2,1 (Netherlands) | No | 2 (3DHistech Pannoramic 250 Flash II), Hamamatsu NanoZoomer XR) | Metastasis detection: AUC=0.895 |
| | **Paige Prostate Detect** | | | | | | | 10,348 WSIs | 1,784 WSIs | 1,1 (USA) | 1,1 (USA) | No | 2 (Leica Aperio AT2; Philips IntelliSite Ultra Fast) | Cancer detection: AUC=0.991; MSK Aperio: AUC=0.991; MSK Philips: AUC=0.964 FNR=0.0174, TPR=0.9826, FPR=0.0500, TNR=0.9500 | 12,727 WSIs, 6,323 patients | [z]800+, 45 | Yes | 1 (Leica Aperio AT2) | Cancer detection: AUC=0.932; FNR=0.0411, TPR=0.9589. FPR=0.1433, TNR=0.8567 |
| **Paige.AI** | [aa]**Paige Prostate Detect** | Novel artificial intelligence system increases the detection of prostate cancer in whole slide images of core needle biopsies[15] | https://doi.org/10.1038/s41379-020-0551-y | May-20 | No | Yes | Internal | NA | Main validation dataset: 232 WSIs<br>Separate validation dataset: 1,811 WSIs | NA | [bb]Unknown, unknown | Un-sure | 1 (Leica Aperio AT2) | Main validation dataset:<br>WSI-level performance: Standalone performance: cancer detection: AUC=0.99, sens=0.96, spec=0.98. Change in pathologist performance (without vs with AI): cancer detection: sens=0.738 vs 0.900; spec: 0.966 vs 0.952; average review time per WSI: 63±39s vs 55±43s.<br>Separate validation dataset:<br>Performance of earlier version of Paige Prostate Alpha: sens=0.98, spec=0.95. | NA | NA | NA | NA | NA |
| **Paige.AI** | [cc]**Paige Prostate Detect** | An independent assessment of an artificial intelligence system for prostate cancer detection shows strong diagnostic accuracy[16] | https://doi.org/10.1038/s41379-021-00794-x | Mar-21 | No | Yes | External | NA | NA | NA | NA | NA | NA | NA | [dd]1,876 WSIs, 118 patients | 1,1 (USA) | No | 1 (Leica Aperio AT2) | Cancer detection: sens=0.977, spec=0.993, PPV=0.979, NPV=0.992, $F_1$=0.98. |
| **Paige.AI** | [ee]**Paige Prostate Detect** | Independent real-world application of a clinical-grade automated prostate cancer detection system[17] | https://doi.org/10.1002/path.5662 | Apr-21 | No | Yes | External | NA | NA | NA | NA | NA | NA | NA | [ff]661 WSIs, 100 patients | 1,1 (Brazil) | No | 1 (Leica Aperio AT2) | Cancer detection (benign vs suspicious):<br>Part-specimen level performance: sens=0.989, spec=0.933, NPV=0.995, PPV=0.865; Patient-level performance: sens=1.00, spec=0.780, NPV=1.00, PPV=0.820; WSI-level performance: AUC=0.996.<br>Change in pathologist performance (without vs with AI): Part-specimen level: sens=0.937 vs 0.966, spec=0.998 vs 0.978, PPV=0.994 vs 0.949, NPV=0.973 vs 0.985; Patient-level: sens=0.940 vs 0.960, spec=0.980 vs 0.920, PPV=0.979 vs 0.923, NPV=0.942 vs 0.958. |

[x] Dataset used for development of three models described in the publication included data from over 800 institutions across 45 countries, and scanned at Memorial Sloan Kettering (MSK) Cancer Centre: USA, UK, Canada, China, Israel, Saudi Arabia, Mexico, Montenegro, Turkey, Brazil, India, Argentina, Australia, Chile, Dominican Republic, Ireland, Pakistan, Peru, Philippines, France, Germany, United Arab Emirates, Ecuador, El Salvador, Greece, Barbados, Croatia, Iceland, Morocco, Poland, Russia, South Africa, Spain, Sri Lanka, Taiwan, Austria, Bangladesh, Belgium, Colombia, Denmark, Lebanon, Norway, Singapore, South Korea, Switzerland. Uncertain whether all labs/countries were represented in the training/test dataset for the breast lymph node model.

[y] External validation test dataset details obtained from CAMELYON16 publication: Ehteshami Bejnordi *et al.*, 2017[14].

[z] Dataset used for development of three models described in the publication included data from over 800 institutions across 45 countries, and scanned at Memorial Sloan Kettering (MSK) Cancer Centre: USA, UK, Canada, China, Israel, Saudi Arabia, Mexico, Montenegro, Turkey, Brazil, India, Argentina, Australia, Chile, Dominican Republic, Ireland, Pakistan, Peru, Philippines, France, Germany, United Arab Emirates, Ecuador, El Salvador, Greece, Barbados, Croatia, Iceland, Morocco, Poland, Russia, South Africa, Spain, Sri Lanka, Taiwan, Austria, Bangladesh, Belgium, Colombia, Denmark, Lebanon, Norway, Singapore, South Korea, Switzerland. Uncertain whether all labs/countries were represented in the external validation dataset for the prostate model.

[aa] Publication describes testing of Paige Prostate Alpha, which is based on the algorithm introduced in Campanella *et al.*, 2019[13].

[bb] Source of data not stated in publication but assumed to be from Memorial Sloan Kettering (MSK) based on author affiliations. Classed as internal validation as MSK was also the original source of model training data.

[cc] Publication notes that: "*The version of software used in this study differs from the originally described version (Raciti et al., 2020) in efficiency and design of the underlying software; the categorization algorithm produces identical results to the original version*".

[dd] Only 1,857 WSIs used for final performance metrics.

[ee] Publication notes that: "*Paige Prostate 1.0 is a convolutional neural network* (*CNN) based on the multiple instance learning algorithm presented in Campanella et al., 2019*[13]*. An early version ('Paige Prostate Alpha') was described in Raciti et al., 2020*[15]*, but the system used and described here is a later version".*

[ff] External validation dataset equates to 579 prostate core needle biopsy 'part-specimens', which is described as the level at which pathology reports are rendered in practice.

| General Details | | | | | | | | Training and Internal Validation | | | | | | | External Validation | | | | |
|---|---|---|---|---|---|---|---|---|---|---|---|---|---|---|---|---|---|---|---|
| **Company name** | **Product name** | **Paper title** | **DOI** | **Date** | **Vendor independence** | **Open access** | **Internal/ external validation** | **Training set:** no. WSIs, cases, patients | **Test set:** no. WSIs, cases, patients | **Training set:** no. data sources, countries | **Test set**: no. data sources, countries | **UK data** | **Test set**: scanner platforms | **Performance** | **Test set:** no. WSIs, cases, patients | **Test set:** no. data sources, countries | **UK data** | **Test set:** scanner platforms | **Performance** |
| **Paige.AI** | [gg]**Paige Prostate Detect** | Clinical Validation of Artificial Intelligence–Augmented Pathology Diagnosis Demonstrates Significant Gains in Diagnostic Accuracy in Prostate Cancer Detection[18] | https://doi.org/10.5858/arpa.2022-0066-OA | Dec-22 | No | Yes | External | NA | NA | NA | NA | NA | NA | NA | [hh]610 WSIs | 218, 1 (USA + others unknown) | Un-sure | 1 (Philips IntelliSite Ultra Fast Scanner Series 2) | WSI-level performance: Cancer detection (benign vs suspicious): AUC=0.99, sens=0.974, spec=0.948. Change in pathologist performance (without vs with AI): sens: 0.887 vs 0.966, spec: 0.973 vs 0.980. |
| **Paige.AI** | **Paige Prostate Detect** | Artificial intelligence–assisted cancer diagnosis improves the efficiency of pathologists in prostatic biopsies[19] | https://doi.org/10.1007/s00428-023-03518-5 | Feb-23 | Yes | Yes | External | NA | NA | NA | NA | NA | NA | NA | 105 WSIs, 41 patients | 1,1 (Portugal) | No | 1 (3DHistech Pannoramic 1000 Dx) | Cancer detection (benign vs suspicious): Pathologist performance (without vs with AI): acc=0.9500 vs 0.9381, sens=0.968 vs 0.955, spec=0.939 vs 0.928, PPV=0.909 vs 0.892; NPV=0.982 vs 0.974; intraobserver concordance=98.81%, κ=0.975. Workflow measures (without vs with AI): IHC requests: 45.95% vs 36.43%; second opinion requests: 12.14% vs 7.38%; median reporting time: 139s vs 108.5s. |
| | **Paige Prostate Grade & Quantify** | | | | | | | | | | | | | | 105 WSIs, 41 patients | 1,1 (Portugal) | No | 1 (3DHistech Pannoramic 1000 Dx) | Grade group prediction: intraobserver concordance = 73.94%, quadratic weighted κ=0.868. |
| [t]***Panakeia Technologies*** | ***PANProfiler Breast*** | *Evaluation of a predictive method for the H&E-based molecular profiling of breast cancer with deep learning*[20] | *https://doi.org/10.1101/2022.01.04.474882* | *Jan-22* | *No* | *Yes* | *Both* | *1,761 WSIs (estimate based on % split, i.e., 85% for training & model selection): 660(ER), 594(PR), 507(HER2)* | *312 WSIs (estimate based on % split, i.e., 15% for testing): 117(ER), 105(PR), 90(HER2)* | [ii]*unknown, unknown (USA + possibly others)* | [ii]*unknown, unknown (USA + possibly others)* | *Un-sure* | *Unknown* | *Biomarker status prediction: acc=0.94 (ER), 0.90 (PR) and 0.89 (HER2); PPV=0.95 (ER), 1.00 (PR), 1.00 (HER2); NPV=0.83 (ER), 0.88 (PR), 0.89 (HER2); Test Replacement Rate*[jj] *(TRR)=0.78 (ER), 0.11 (PR), 0.52 (HER2).* | [kk]*Model calibration=809 WSIs: 278(ER), 267(PR), and 264(HER2) Test set=1,047 WSIs: 370(ER), 381(PR), and 296(HER2);* | *23 (estimated), 1 (USA)* | *No* | *unknown* | *Biomarker status prediction: acc=0.87 (ER), 0.83 (PR), 0.87 (HER2), 0.90 (HER2-ISH*[ll]*). TRR=0.89 (ER), 0.48 (PR), 0.78 (HER2), 0.82 (HER2-ISH); PPV=0.87 (ER), 0.83 (PR); NPV=0.86 (ER), 0.79 (PR), 0.87 (HER2), 0.90 (HER2-ISH)* |
| **Primaa** | **Cleo Breast** | Multicenter automatic detection of invasive carcinoma on breast whole slide images[21] | https://doi.org/10.1371/journal.pdig.0000091 | Feb-23 | No | Yes | Both | 952 WSIs, 148 patients | 88 WSIs, 25 patients | [mm]1, 1 (assumed) (France) | [mm]unknown | Un-sure | 1 (Hamamatsu S120) | WSI-level performance: Cancer detection: accuracy=0.976, precision=1.00, recall=0.909 | Fine tuning/calibration: 72 WSIs, 18 patients Test set: 83 WSIs, 14 patients | [mm]1,1 (assumed) (France) | Un-sure | 1 (3DHistech P1000) | WSI-level performance: Cancer detection: acc=0.920, precision=0.708, recall=1.00. |
| [t]***Primaa*** | ***Cleo Breast*** | *Automatic detection of microcalcifications in whole slide image - comparison of deep learning and standard computer vision approaches*[22] | https://dx.doi.org/10.36227/techrxiv.21981614.v2 | *Feb-23* | *No* | *Yes* | *Both* | *1442 WSIs* | *24 WSIs* | *1, 1 (unknown)* | *1, 1 (unknown)* | *Un-sure* | *1 (Hamamatsu)* | *Slide-level metrics:* [nn]*Acc_top16 = 0.81, FP_top16=6.2. Patch-level metrics: AUC=0.985, acc=0.99, recall=0.8, precision=0.53* | *95 WSIs* | *3, unknown (unknown)* | *Un-sure* | *2 (Hamamatsu and 3DHistech)* | *Performance across three centres providing test data: Slide-level metrics:* [nn]*Acc_top16 = 0.77-94, FP_top16=6.9-12.8. Patch-level metrics: Acc=0.92-99, recall=0.46-82, precision=0.05-0.57* |

[gg] Publication describes Paige Prostate, which is a more mature version of Paige Prostate Alpha.

[hh] External validation dataset includes data from Memorial Sloan Kettering (MSK), which is the same institution from which data was used for model training. Data was gathered from 217 other institutions of unspecified location.

[ii] Training/internal validation datasets included data from the TCGA-BRCA dataset and data from a private clinical data provider (BioIVT). Unclear how many TCGA labs were included in the training/internal validation and number of sources/countries represented in BioIVT dataset is unknown.

[jj] Test replacement rate (TRR): described as a measure of the effectiveness of the device to return confident predictions, which corresponds to the % of predictions performed over all classes. Estimates the number of traditional tests that could be replaced by PANProfiler Breast for molecular profiling.

[kk] External validation dataset was comprised of the TCGA-BRCA dataset, but from different labs to those used in training/internal validation. Figure 5 suggests at least 23 of 26 labs were represented in the dataset.

[ll] HER2-ISH refers to concordance of the algorithm results with *in situ* hybridisation (ISH) tests.

[mm] Dataset source unknown. Data availability statement suggests at least some data is from a French tissue bank. The publication describes training on a dataset from a '*reference acquisition center*' and testing on a *'new target acquisition center'*.

[nn] 'acc_top16' metric: will be 1 if, among the first 16 patches, a true microcalcification is present; else 0. 'FP_top16' metric: number of false positives (FPs) among the first 16 patches.

| General Details | | | | | | | | Training and Internal Validation | | | | | | | External Validation | | | | |
|---|---|---|---|---|---|---|---|---|---|---|---|---|---|---|---|---|---|---|---|
| **Company name** | **Product name** | **Paper title** | **DOI** | **Date** | **Vendor independ-ence** | **Open access** | **Internal/ external validation** | **Training set:** no. WSIs, cases, patients | **Test set:** no. WSIs, cases, patients | **Training set:** no. data sources, countries | **Test set**: no. data sources, countries | **UK data** | **Test set**: scanner platforms | **Performance** | **Test set:** no. WSIs, cases, patients | **Test set:** no. data sources, countries | **UK data** | **Test set:** scanner platforms | **Performance** |
| **Stratipath** | [oo]**Stratipath Breast** | Improved breast cancer histological grading using deep learning[23] | https://doi.org/10.1016/j.annonc.2021.09.007 | Sep-21 | No | Yes | Both | 844 WSIs/patients | Classification performance (NHG1&3 cases): 351 WSIs/patients.<br><br>Prognostic performance (NHG2 cases): 372 WSIs/patients. | [pp]3+, 2 (Sweden, USA) | 3+, 2 (Sweden, USA) | No | 2 (Hamamatsu Nanozoomer XR and Hamamatsu Nanozoomer S360) | Invasive tumour detection (benign vs cancer): weighted acc=0.87, $F_1$=0.85, precision=0.91, recall=0.80.<br><br>WSI-level performance:<br>[qq]classification (NHG1 vs 3): weighted acc=0.84-0.89, AUC=0.919-0.937, $F_1$=0.85-0.92, precision=0.86-0.98, recall=0.82-1.<br><br>Prognostic performance (risk of recurrance in stratified NHG2 DG2-high vs DG2-low patients): HR=2.94 | Classification performance (NHG1&3 cases): 654 WSIs/patients;<br><br>Prognostic performance (NHG2 cases): 608 WSIs/patients | 1,1 (Sweden) | No | Unknown | WSI-level performance:<br>classification (NHG1 vs 3): weighted acc=0.83, AUC=0.907, $F_1$ score=0.87, precision=0.83, recall=0.85,<br>Prognostic performance (risk of recurrence in stratified NHG2 DG2-high vs DG2-low patients): HR=1.91 |
| **Visiopharm** | **Metastasis Detection, AI** | Artificial Intelligence-Aided Diagnosis of Breast Cancer Lymph Node Metastasis on Histologic Slides in a Digital Workflow[24] | https://doi.org/10.1016/j.modpat.2023.100216 | May-23 | Yes | No | External | NA | NA | NA | NA | NA | NA | NA | Sentinel lymph node (SLN) validation cohort: 234 LNs<br><br>SLN consensus cohort: 102 LNs<br><br>Non-sentinel LN (NSLN) validation cohort: 258 LNs | [rr]1,1 (USA) | No | 1 (Philips IntelliSite Ultra Fast Scanner) | SLN metastasis detection (validation cohort): sens=1.00, spec=0.415, PPV=0.295, NPV= 1.00. Concordance with pathologists (consensus cohort): 98-99%.<br>NSLN metastasis detection (validation cohort): sens=1.00, spec=0.785, PPV=0.681, NPV=1.00. |

[oo] The final product is for stratification of NHG2 patients only.

[pp] Internal validation dataset included data from two hospitals in Sweden and from TCGA (collected from multiple USA labs). Exact number of labs and scanner for TCGA dataset unknown.

[qq] Internal validation performance is presented for each test set from each data source separately, hence a range of values is displayed.

[rr] Data source unknown, but publication states that slides were scanned "*as a part of the digital workflow during our routine pathology practice*" so data assumed to be from Ohio State University, USA, based on author affiliations.

#### Glossary & Abbreviations

**Accuracy (Acc):** TP + TN / TP+FP+TN+FN (True Positive + True Negative/True Positive + False Positive + True Negative + False Negative) i.e., the number of correct predictions, as a proportion of all samples.

**Sensitivity (Sens):** TP / TP+FN (True Positive/True Positive + False Negative) i.e., the number of samples correctly predicted as positive, as a proportion of all positive samples.

**Specificity (Spec):** TN / TN + FP (True Negative/True Negative + False Positive) i.e., the number of samples correctly predicted as negative, as a proportion of all negative samples.

**Precision:** TP / TP+FP (True Positive/True Positive + False Positive) i.e., number of samples correctly predicted as positive, as a proportion of all samples predicted as positive.

**Recall:** *see Sensitivity.*

**True Negative Rate (TNR):** *see Specificity.*

**True Positive Rate (TPR):** *see Sensitivity.*

**False Negative Rate (FNR):** FN / FN + TP (False Negative/False Negative + True Positive): i.e., number of samples incorrectly predicted as negative, as a proportion of all positive samples.

**False Positive Rate (FPR):** FP / FP + TN (False Positive/False Positive + True Negative) i.e., number of samples incorrectly predicted as positive, as a proportion of all negative samples.

**Negative Predictive Value (NPV):** TN / TN + FN (True Negative/True Negative + False Negative) i.e., number of samples correctly predicted as negative, as a proportion of all samples predicted as negative.

**Positive Predictive Value (PPV):** *see Precision.*

**AUC:** Area Under the Receiver Operating Characteristic (ROC) curve. The ROC curve for an algorithm shows the TPR (y-axis) against the FPR (x-axis). The AUC provides a measure of algorithm performance across all potential classification thresholds.

**AUC(PR)**: Area Under the Precision-Recall (PR) curve. The PR curve for an algorithm shows the precision (PPV, y-axis) against the recall (TPR, x-axis). The AUC(PR) is typically used to show algorithm performance in situations where the priority objective is to identify positive samples.

**$F_1$ score ($F_1$):** a measure of accuracy defined as the harmonic mean of precision and recall (2TP / 2TP + FP + FN).

**Cohen's Kappa coefficient (κ):** a measure of concordance between two ratings of a given variable. In this context, it could be between the concordance between the algorithm output and the ground-truth as determined by pathologists, or between two different pathologists, or within the same pathologist on multiple reads.

**Weighted Cohen's Kappa/Quadratic weighted Cohen's Kappa coefficient (weighted κ/quadratic weighted κ):** weighting of Cohen's Kappa to assess concordance between two ratings of an ordinal variable (e.g., Grade Group), which allows different weighting of classification disagreements.

**Matthews Correlation Coefficient (MCC):** a measure of the association between two variables, e.g., the algorithm-predicted classifications and the ground truth as determined by pathologists. It is used to describe the confusion matrix with a single value.

**Pearson Correlation Coefficient (r):** a measure of the linear correlation between two variables, e.g., the algorithm-predicted classifications and the ground truth as determined by pathologists.

**Spearman's Rank Correlation Coefficient (Spearman's rho):** a measure of the correlation between two variables for nonparametric data.

**Harrell's concordance index (C-index):** a measure of concordance for algorithms which produce a risk score as an output (e.g., between the algorithm-predicted prognosis and actual patient survival).

**Hazard Ratio (HR):** an estimate of the relative risk of an event occurring by taking the ratio of the hazard rate between two groups, e.g., patient survival between different prognostic groups.

**WSI**: whole slide image.
